# Functionally Focused Evaluation: A Novel Comparative Protocol for Wearable Electroencephalography Headsets

**DOI:** 10.64898/2026.06.03.26354802

**Authors:** Anand Bhuyan, Michael Wong, Alistair McEwan, Cameron Higgins, Navin Cooray

**Affiliations:** School of Biomedical Engineering, The University of Sydney, Sydney, NSW, Australia; Cerebral Palsy Alliance, Discipline of Child and Adolescent Health, The University of Sydney, Sydney, NSW, Australia; Resonait Medical Technologies Pty Ltd, Sydney, New South Wales, Australia; Commonwealth Scientific and Industrial Research Organisation (CSIRO), Canberra, ACT, Australia

**Keywords:** Electroencephalography (EEG), quality assessment, wearable systems

## Abstract

With the emergence of electroencephalography (EEG) as a tool in the cognitive domain, new demands are being placed on the technology to keep up with functional applications – especially in the context of at-home neural monitoring. New use cases have fostered development of wearable EEG (wEEG) devices: portable, low-cost headsets used for EEG monitoring. This evolution of technology and application has not been accompanied by development in technology evaluation, often relying on function-agnostic markers to assess devices for efficacy in this new space. With current methods limited in scope, this study designed, tested and evaluated a novel functionally-focused comparative protocol for wEEG devices. Eight participants undertook a protocol for the evaluation of four established wEEG devices, assessing cognitive resolution and general usability. Compared to a well-established traditional analysis method (eyes open/eyes closed protocol), the novel design proposed here enabled the same analysis of headset resolution, while also providing additional context into user preferences and opening downstream possibilities for specific cognitive insights. Future research could enable the development of this protocol into a standardised method to ensure the performance of wEEG technology can satisfy emerging clinical needs.

## 1. Introduction

Electroencephalography (EEG) is a neural analysis method involving the non-invasive measurement of the brain’s electrical fields, used to approximate cognitive activity [1]. While traditionally utilized in clinic to further our understanding of the human brain (either on an individual or population level), new applications have emerged, ranging from the diagnosis, treatment and support of mental health conditions [2-4] to brain-computer interface (BCI) control of assistive technology [5, 6]. These emerging clinical applications have led to the translation of EEG technology to at-home use via the design and development of wearable EEG headsets – portable, lower-cost alternatives to the in-clinic technology [7-9]. While these wearable headsets have opened up new, important pathways, the benefits they bring come at a signal quality cost [10]. Combined with the diversity in structure and function of these headsets, with a range of different electrode types and quantities [11], this compromise on quality means headsets need to be evaluated effectively to ensure they are fit for clinical use. In addition to technology selection on a case-by-case basis, effective performance evaluation is needed to enable further research and development in this area, facilitating the design of technology that can better optimise the trade-off between accuracy and accessibility aligned with these functional applications [12].

Typical methods used to comparatively evaluate headsets though are increasingly out-of-date, incapable of completely assuring that wEEG technology can pick up on cognitive activity with the sensitivity and specificity required by these newer applications. These traditional methods of performance evaluation tend to be locally focused, evaluating signal quality at individual electrodes compared to the same electrode in a gold-standard system (and not attempting comparison between different wEEG options) [8, 13]. Other oft-used methods assess resolution using local signal patterns, for example comparing technology on its ability to pick up specific EEG trends like event-related potentials (ERPs) or the difference in EEG signal with eyes open as compared to with eyes closed (the Berger effect) [14, 15]. While these methods may be effective in validating electrode design and even local resolution, they are not necessarily informative on how well a headset might pick up on more complex cognitive patterns that are key to the emerging clinical applications of this technology. As such, a new type of performance evaluation protocol that can assess entire system performance in detecting different types of brain activity is needed. Performance evaluation using cognitive tasks to understand how well headsets can pick up local neural markers have shown promise [16, 17], but have not yet been sufficiently generalisable to the spectrum of potential clinical use cases and still face challenges in mapping connected and distributed neural responses that may be of clinical interest. This raises the question of whether we are able to design a wEEG validation protocol that can both encompass a more connected view of neural activity while still placing focus on functional relevance.

Multi-variate pattern analysis (MVPA) methods are widely used within cognitive neuroscience to assess the differentiability of different types of neural activity using machine learning techniques [18-22]. As imaging studies have shown that cognitive tasks can be chosen to specifically elicit activation in different functional networks throughout the brain, we can therefore apply an MVPA approach to this performance validation problem, analysing how effectively wEEG devices can pick up relative differences in cognitive activity between different cognitive tasks, and therefore by proxy different resting-state networks (RSNs) within the brain.

Beyond the evaluation of function, the move to at-home usage of this technology has meant a shift from use being heavily monitored by a trained professional, to applications where adherence is entirely user-dependent. Overcomplexity of technology can be extremely costly in terms of clinical value [23], especially with use in these settings where user skill can create a technological barrier to reaching the clinical benefits of technology usage [24]. Headset comfort is an influential factor in headset use, especially on a regular basis [25], and yet many wEEG headsets are not inherently user-friendly [26]. As such, the design of a new protocol also necessitates inclusion of some measure of user-friendliness, with functional application immaterial if devices are not likely to be used. Analysis of usability alone has been performed with wEEG technology in the past, assessment is rarely standardised, often creating arbitrary categories that as a consequence of inconsistency, lose relevance across the spectrum of research in this area [25-28]. The novel protocol design proposed here should hence seek to incorporate standardized assessment of technology usability to accompany judgement on the functional capabilities of wEEG devices.

This experiment therefore seeks to fill the gap in performance validation of wEEG headsets by designing a novel MVPA-based hardware evaluation protocol incorporating usability testing and evaluating this protocol in the analysis of four existing wEEG devices. The proposed protocol consists of a series of cognitive tasks, each chosen to elicit activation in specific RSNs of interest, followed by a standardised usability test to assess user-friendliness. wEEG can then be assessed for quality on the strength of how well they enable decoding of different RSN activation, as well as how usable they are – two properties that are integral to modern use cases of this technology. For the purposes of this study, RSNs were chosen for inclusion on the basis of three key factors. Firstly, activation of each RSN should be reliably detectable with EEG technology. Secondly, each RSN must have functional relevance. Finally, each RSN should have evidence of reliable activation with a simple cognitive task. On the basis of these three factors, four networks were chosen including the Default Mode Network (DMN) [29-36], the Dorsal Attention Network (DAN) [37-43], the Linguistic Network (LN) [44-47], and Sensorimotor Network (SMN) [48-52]. The four cognitive tasks included in this study therefore were chosen with respect to these four networks. Evaluation of the novel design was completed by putting eight participants through this protocol and comparing measured headset quality differences with respect to a current method of technology comparison. As there is a significant level of inconsistency in existing performance evaluation methods, a well-established method generally used for this type of analysis was chosen for baseline comparison – the ability of devices to detect the Berger effect [15] i.e. how well headsets can discriminate between eyes closed and eyes open EEG signal [53-58].

We hypothesise that use of this novel protocol design across a small participant group will support headset differences identified using traditional methods, while also providing insight into user preferences and ensuring headset analysis is rooted in specific cognitive resolution.

## 2. Method

### 2.1. Experimental Design

Eight participants completed the experiment, where each participant provided two scans with four different wEEG headsets, providing eight scans in total. The device manufacturer has been anonymised to emphasise device characteristics rather than specific brands (technical specifications and characteristics are provided in the Table 1).

**Table 1.** Characteristics of the four headsets analysed here.

| Device | Electrode Quantity | Electrode Type |
| --- | --- | --- |
| Device 1 | 5 | Gel |
| Device 2 | 2 | In-Ear |
| Device 3 | 32 | Saline |
| Device 4 | 16 | Saline |

One scan with any given headset involved the participant completing a series of 36 paradigms, each with a 30-second duration. The first and last six paradigms within each scan were not a part of the novel protocol design but were included as a baseline or control phase representing traditional, univariate methods of comparison. This baseline phase consisted of alternating 30-second instances of eyes open, within which participants were asked to fixate on a white cross on a blank black screen, and eyes closed, within which participants were merely asked to close their eyes until otherwise instructed. The middle 24 paradigms represented the new protocol design, consisting of four cognitive tasks appearing six times each, all chosen to elicit specific functional activation in a clinically relevant RSN. The cognitive tasks were presented in six blocks of four, with each task appearing exactly once in each block, but in randomised order. The cognitive tasks in question included an autobiographical memory task for DMN activation, an auditory oddball task for DAN activation, a listening comprehension task for LN activation and a motor imagery task for SMN activation.

Completing the autobiographical memory task entailed reflecting on a clear childhood memory for the duration of the 30-second task instance. Participants were briefed to reflect on the same memory each time they completed this task and to do so with their eyes open, fixating on a white cross on the screen. This task was chosen to elicit DMN activation, with DMN activity consistently linked to memory [59, 60].

Performance of the auditory oddball task required participants to listen to a series of 60 tones (of 0.25 second duration) played across the 30-second period while fixating on the white cross with their eyes open. They were asked to track the number of oddball tones (600 Hz) delivered interspersed randomly with normal tones (440 Hz) throughout the duration of each 30-second task instance. The probability of any given being an oddball tone was 0.14. This task was designed to elicit DAN activation [61, 62].

Completion of the motor imagery task asked participants to mentally rehearse the action of squeezing a stress ball for the duration of the 30-second task instance, while fixating on the white cross with eyes open. Prior to completing every scan, participants were given a stress ball to physically experience performing the action, ensuring they would be able to mentally rehearse it accurately. This task was chosen to elicit SMN activation, with motor imagery tasks continually shown to be sufficient to impact activity throughout motor areas within the brain [63, 64].

The listening comprehension task needed participants to listen to a 30-second snippet of an open-source English audiobook version of a short story from Hans Christian Andersen’s Fairy Tale Collection, entitled the Emperor’s New Suit [65], while fixating on the white cross with eyes open. Each 30-second snippet played for participants would be different both within and between scans. This task was designed to activate the LN within the brain, with activity in these areas shown to be elicited by listening to speech [66-68].

Throughout the completion of all of cognitive tasks, participants were also asked to fixate on a white cross on an otherwise blank black screen. The experimental design in full is detailed in Figure 1.

**Figure 1.**
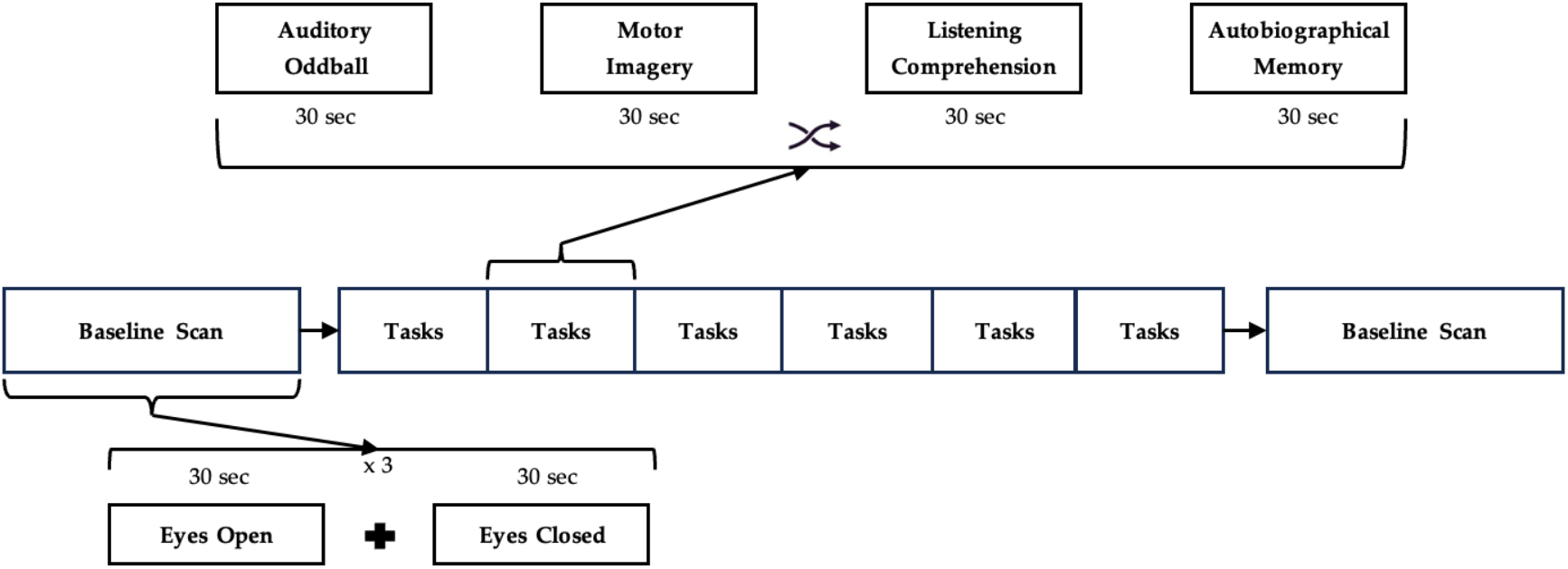
Experimental design.

All participants were given sufficient instruction prior to every session on how to complete every task correctly. Every scan consisted of the same 36 tasks, with alternating eyes open and eyes closed tasks to begin and end the session, but the order of the cognitive tasks in between was randomised to eliminate any potential biasing. The order of headsets to perform ‘scans’ with was also randomised, using a Latin Square model to ensure further bias avoidance [69], with all participants performing one scan with each headset before commencing their second round of scans (Table 2).

**Table 2.** Participant scan order via randomised Latin Square.

| Participant # | Scan Order |  |  |  |  |  |  |  |
| --- | --- | --- | --- | --- | --- | --- | --- | --- |
|  | 1 | 2 | 3 | 4 | 5 | 6 | 7 | 8 |
| 1 | Device 1 | Device 2 | Device 4 | Device 3 | Device 3 | Device 4 | Device 2 | Device 1 |
| 2 | Device 1 | Device 2 | Device 4 | Device 3 | Device 3 | Device 4 | Device 2 | Device 1 |
| 3 | Device 2 | Device 1 | Device 3 | Device 4 | Device 4 | Device 3 | Device 1 | Device 2 |
| 4 | Device 2 | Device 1 | Device 3 | Device 4 | Device 4 | Device 3 | Device 1 | Device 2 |
| 5 | Device 3 | Device 4 | Device 2 | Device 1 | Device 1 | Device 2 | Device 4 | Device 3 |
| 6 | Device 3 | Device 4 | Device 2 | Device 1 | Device 1 | Device 2 | Device 4 | Device 3 |
| 7 | Device 4 | Device 3 | Device 1 | Device 2 | Device 2 | Device 1 | Device 3 | Device 4 |
| 8 | Device 4 | Device 3 | Device 1 | Device 2 | Device 2 | Device 1 | Device 3 | Device 4 |

After completing the second scanning session with each device, participants were asked to complete a standardised usability survey following the System Usability Survey (SUS) model to assess user-friendliness [70]. This data was then completely anonymised and deidentified before handover for the purpose of analysis here. The experimental data was provided by Resonait Medical Technologies and comprised of 36 Comma-Separated Values (CSV) files of EEG data and 36 text files (containing labels defining the task being concurrently performed) for each scan completed, with 64 scans completed in total – 16 scans with each of the four headsets, with two completed by each of the eight participants for each headset. In addition, a CSV file of usability survey responses was also passed on, with eight sets of responses (one per participant) for each headset, collected anonymously using Google Forms. This study received ethics approval under The University of Sydney Research Integrity and Ethics Administration (2024/HE001864).

### 2.2. Participant Demographic

Eight participants completed the protocol described, with five male and three female. Mean age across the eight participants was 27.13 (Table 3). Participant recruitment was performed entirely by Resonait Medical Technologies and selection was not necessarily in line with typical expected end-users for these devices nor representative of a broad cross-section of society. All demographic information recorded in Table 1 was supplied by Resonait Medical Technologies.

**Table 3.** Demographic information of study participants.

| Category |  | Number of Participants |
| --- | --- | --- |
| Sex | Female | 3 |
|  | Male | 5 |
|  | Other | 0 |
| Age | Mean | 27.13 |
|  | Standard Error (SE) | 2.61 |

### 2.3 Data pre-processing

Data was extracted scan by scan, iterating through the 36 data files for each scan corresponding to the 36 30-second task instances completed, extracting the EEG signals into one data matrix and storing respective task labels for each sample. The EEG from all headsets was sampled at 128 Hz. For each scan, all dropouts were removed, the data was demeaned, slow drifts were removed via 5^th^ order polynomial detrending applied to each channel, then passed through a fifth-order zero phase Butterworth filter for band-pass filtering to remove high-frequency noise (using MATLAB’s inbuilt ‘butter’, ‘detrend’ and ‘filtfilt’ functions). The first and last 0.5seconds were cropped to remove any filter artefacts, with the data then normalised over each channel.

After data filtering, within each scan, all the EEG data was segmented into 0.25-second epochs for feature extraction – with each 0.25-second epoch containing 32 samples. For any given scan, this yielded 4032 epochs, with 672 attributed to each task label (112 0.25-second epochs from each 30-second task after data pre-processing). For each epoch, a lagged covariance matrix was computed as a feature representation. Given an epoch data matrix X ∈ ℝ^(T×N), we formed the augmented matrix [X, X_(−τ)] ∈ ℝ^(T×2N) by horizontally concatenating X with a lagged copy of itself at τ = 5 samples and computed the (2N × 2N) sample covariance matrix of this augmented matrix. As such, the total feature set length was variable, depending on the number of headset channels (which was inconsistent between headsets). These features were then aggregated across all epochs to give a matrix with 4032 rows of features (one for each of the 4032 epochs).

## 2.4. Data Analysis

### 2.4.1. Baseline Condition

As a baseline condition, we evaluated binary classification of eyes-open (EO) versus eyes-closed (EC) EEG segments. For each scan, epochs from five of six 30-second blocks per condition were used to train a linear SVM classifier (MATLAB’s fitclinear, L2 regularisation), with the remaining block from each condition held out for testing. Class labels for held-out epochs were predicted individually, and a final classification decision was made by majority vote across epochs within each held-out block. Classification accuracy was computed as the proportion of correctly classified held-out blocks across the six folds.

Leave-one-out cross-validation was performed to ensure no bias was introduced by data splitting, testing on the first instance of the tasks, then the second instance and so on. Overall classification accuracy for each scan was calculated by averaging the classification accuracy across the six cross-validation folds, giving a single classification accuracy output for each of the 16 scans performed for all four headsets.

While traditional ML methods use a variety of other performance metrics to quantify classifier performance (e.g. sensitivity, specificity, precision, F1 score [73]), these are usually designed to understand performance in dealing with specifically ‘positive’ or specifically ‘negative’ cases. While these metrics are relevant in classification problems where one condition is diseased and one is healthy, they are not significant in this work, within which the two conditions are different, but neither is reflective of disease or health. As such, accuracy was chosen as the metric of classifier performance and therefore differentiability of the compared signals.

To enable headset comparison, the accuracies obtained for each headset were averaged across the 16 scans, giving one classification accuracy per headset. Additionally, paired t-test plots were created for the 16 classification accuracies for each headset, separated by participants’ first and second scans, to illustrate uniformity or lack thereof.

#### 2.4.2. Cognitive Comparisons

For the experimental, ‘novel’ condition, consisting of assessing the differentiation between the four cognitive tasks incorporated within the novel protocol design (referred to interchangeably from here onwards as ‘cognitive comparisons’), features extracted from the EEG signal recorded during the four cognitive tasks within each scan were analysed. For every scan, the four cognitive tasks were compared in pairs, representing the comparison of EEG signals theoretically linked with activation of two different RSNs, with four tasks meaning six pair-wise comparisons (autobiographical memory versus auditory oddball, autobiographical memory versus motor imagery, auditory oddball versus motor imagery, autobiographical memory versus listening comprehension, auditory oddball versus listening comprehension and motor imagery versus listening comprehension).

For each pairwise comparison, epochs from five of six 30-second blocks per task were used to train a linear SVM classifier (MATLAB’s fitclinear, L2 regularisation), with the remaining block from each task held out for testing. Class labels for held-out epochs were predicted individually, and a final classification decision was made by majority vote across epochs within each held-out block. Classification accuracy was computed as the proportion of correctly classified held-out blocks across the six folds.

Once again, leave-one-out cross-validation was performed to ensure no bias was introduced by data splitting, and overall classification accuracy for each cognitive comparison within each scan was calculated by averaging the classification accuracy across the six cross-validation folds for a given comparison.

Classification accuracy was averaged across the six cognitive comparisons within each scan, giving a ‘mean cognitive comparison’ classification accuracy for each of the 16 scans performed with each of the four headsets. As seen in 2.4.1., the mean cognitive comparison accuracies were averaged across the 16 scans for each headset, giving one classification accuracy per headset. Again, paired t-test plots were used to visualise the relationship between participants’ first and second scans by headset.

### 2.4.3. Usability Analysis

SUS survey response values vary from one to five, where one represents ‘Strongly Disagree’ and five represents ‘Strongly Agree’ with the survey question, which were analysed according to the SUS model. As per SUS convention [70], a total SUS usability score can be calculated from the response values given. This analysis was all performed in Microsoft Excel, before being exported into MATLAB for statistical analysis.

### 2.5. Statistical Analysis

All graphing and statistical analysis was performed using MATLAB for this project. All means and standard errors across both experimental conditions were also calculated using MATLAB.

#### 2.5.1. Linear Mixed Effects Models and Analysis of Variance (ANOVA)

Eyes Open/Eyes Closed condition classification accuracies and mean cognitive comparison classification accuracies were both analysed using a Linear Mixed Effects model [74] (via MATLAB’s ‘fitlme’ function), allowing for fixed participant effects from the same participants completing the experiment with all four devices. On this Linear Mixed Effects model, an Analysis of Variance (ANOVA), using MATLAB’s inbuilt ‘anova’ function, was performed between the sets of classification accuracies for each device to determine whether the device type (a variable effect) had a meaningful impact on classification accuracy. If device type was judged to have a significant effect (p < 0.05), further analysis was then performed via pair-wise device by device comparisons, using coefficient tests (through MATLAB’s inbuilt ‘coefTest’). This enabled analysis of statistical differences between every device for both conditions, allowing for directional comments about device resolution.

Analysis of usability outcomes was also performed using this model, but the participant was not assessed as a fixed effect as anonymised responses made it impossible to link responses across the four headsets for each participant. Therefore, instead a normal linear model was used (via MATLAB’s ‘fitlm’). As above, an ANOVA was performed across the linear model to evaluate if device type had an effect on classification accuracy. Again, if accuracy was found to vary over device types, coefficient tests were performed to compare devices in pairs and identify individual differences.

## 3. Results

### 3.1 Comparing Headsets Using Traditional Methods

#### 3.1.1 Variations between Headsets

Analysing the differentiation of Eyes Open/Eyes Closed EEG signals, showed Device 1 to distinguish between these signals with a mean classification accuracy of 0.84, Device 2 with an accuracy of 0.54, Device 3 with an accuracy of 0.98 and Device 4 with an accuracy of 0.89 (see Figure 2). Device type was found to have a significant effect on classification accuracy (p < 0.001). Given the significance of device type, pair-wise comparisons were performed between each device. Device 3 facilitated higher classification accuracy than Device 1 (p = 0.004) and Device 2 (p < 0.001). Device 1 saw significantly better classification accuracy than Device 2 (p < 0.001), as did Device 4 (p < 0.001). Device 4 and Device 1 did not differ meaningfully (p = 0.287), nor did Device 4 and Device 3 (p = 0.065).

**Figure 2.**
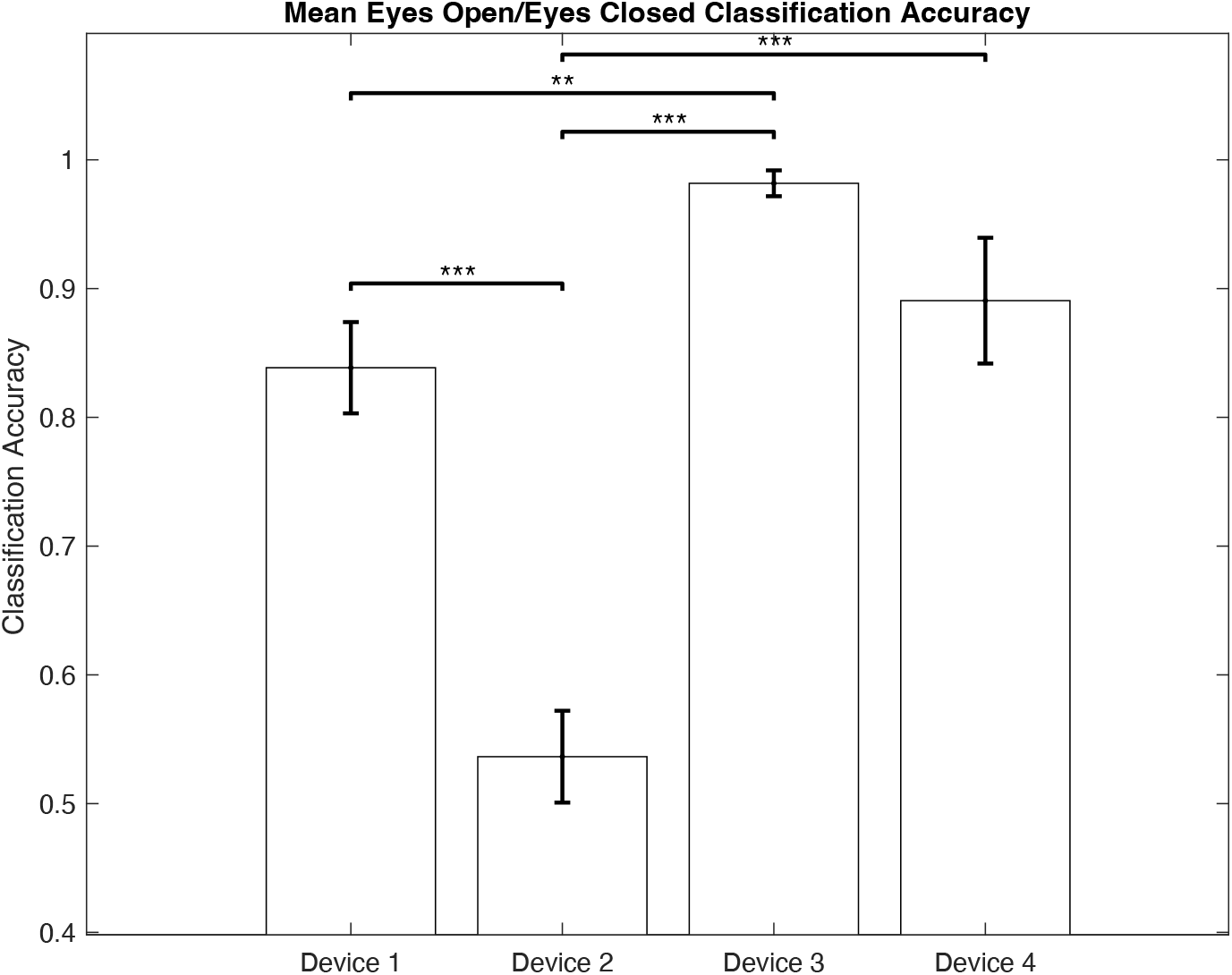
SVM classification accuracy of Eyes Open/Eyes Closed EEG data averaged across scans. Error bars display standard error, and brackets display statistical difference as follows: * - p < 0.05, ** - p < 0.01, *** - p < 0.001.

#### 3.1.2 Variations within and between Participants

Classification accuracy between Eyes Open/Eyes Closed signal varied both between participants and within participants, as demonstrated in Figure 3. Classification accuracies did not differ from participants’ first scan and second scan for Device 1 (p = 0.686), Device 2 (p = 0.634), Device 3 (p = 0.088) or Device 4 (p = 0.283).

**Figure 3.**
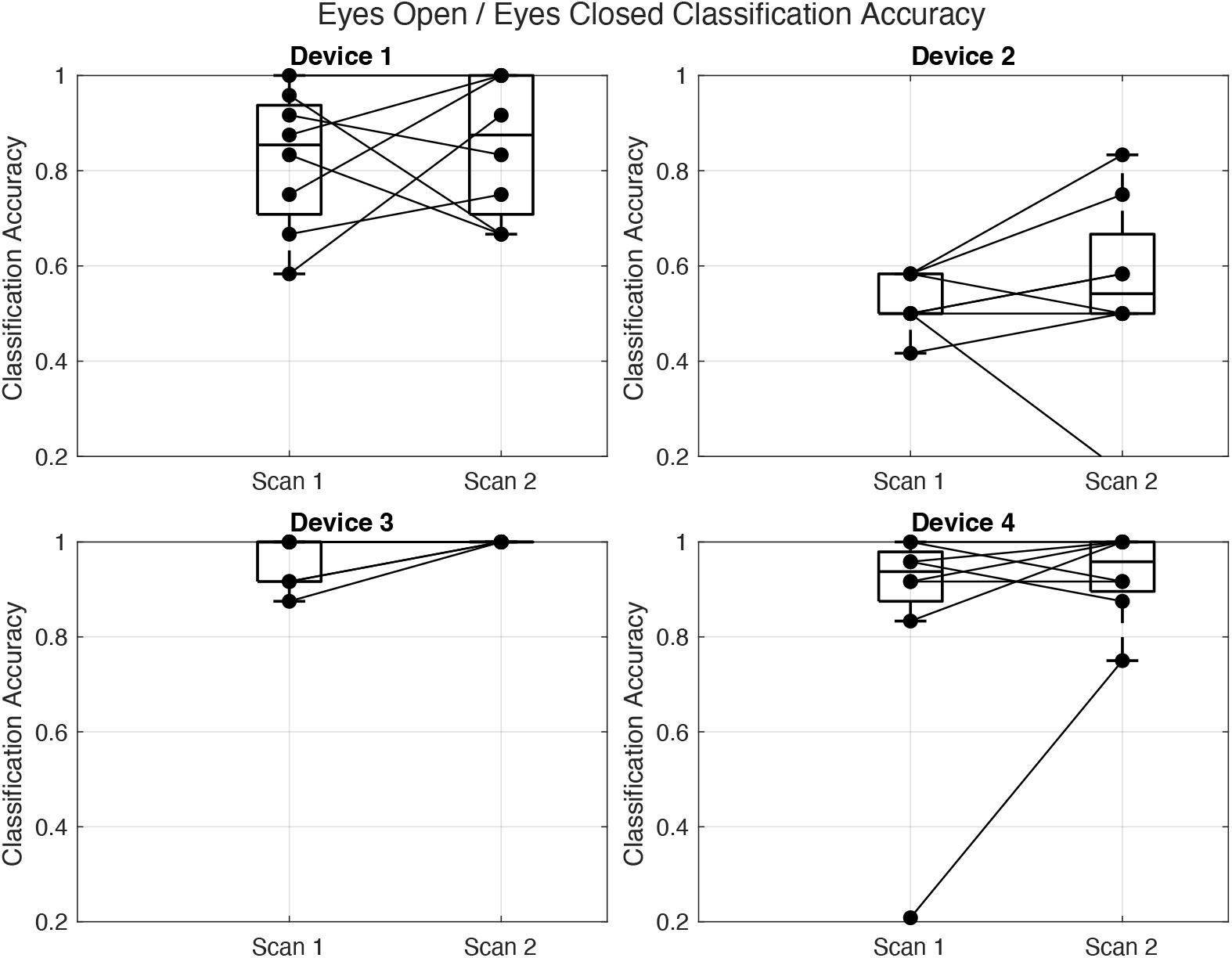
Paired t-test plot of SVM classification accuracy for Eyes Open/Eyes Closed EEG data by device. Accuracies have been separated out by scan, with classification accuracies corresponding to the same participant linked for each device.

### 3.2 Comparing Headsets Using Novel Comparative Protocol

#### 3.2.1 Variations between Headsets

Analysing mean classification accuracy across headsets for pairwise cognitive comparisons, i.e. how well tasks and therefore RSN activation could be decoded for each headset, showed Device 1 was able to differentiate between cognitive tasks at an accuracy of 0.56, Device 2 at an accuracy of 0.50, Device 3 at an accuracy of 0.62 and Device 4 at an accuracy of 0.58 (see Figure 4). Again, device was a meaningful effect on classification accuracy (p < 0.001), justifying further analysis, comparing headsets pairwise. Device 2 was found to experience lower classification accuracy than Device 1 (p = 0.019), Device 3 (p < 0.001) and Device 4 (p = 0.004). Device 3 outperformed Device 1 (p = 0.023). Device 4 and Device 1 did not meaningfully differ in accuracy across these cognitive tasks (p = 0.555), nor did Device 4 and Device 3 (p = 0.088).

**Figure 4.**
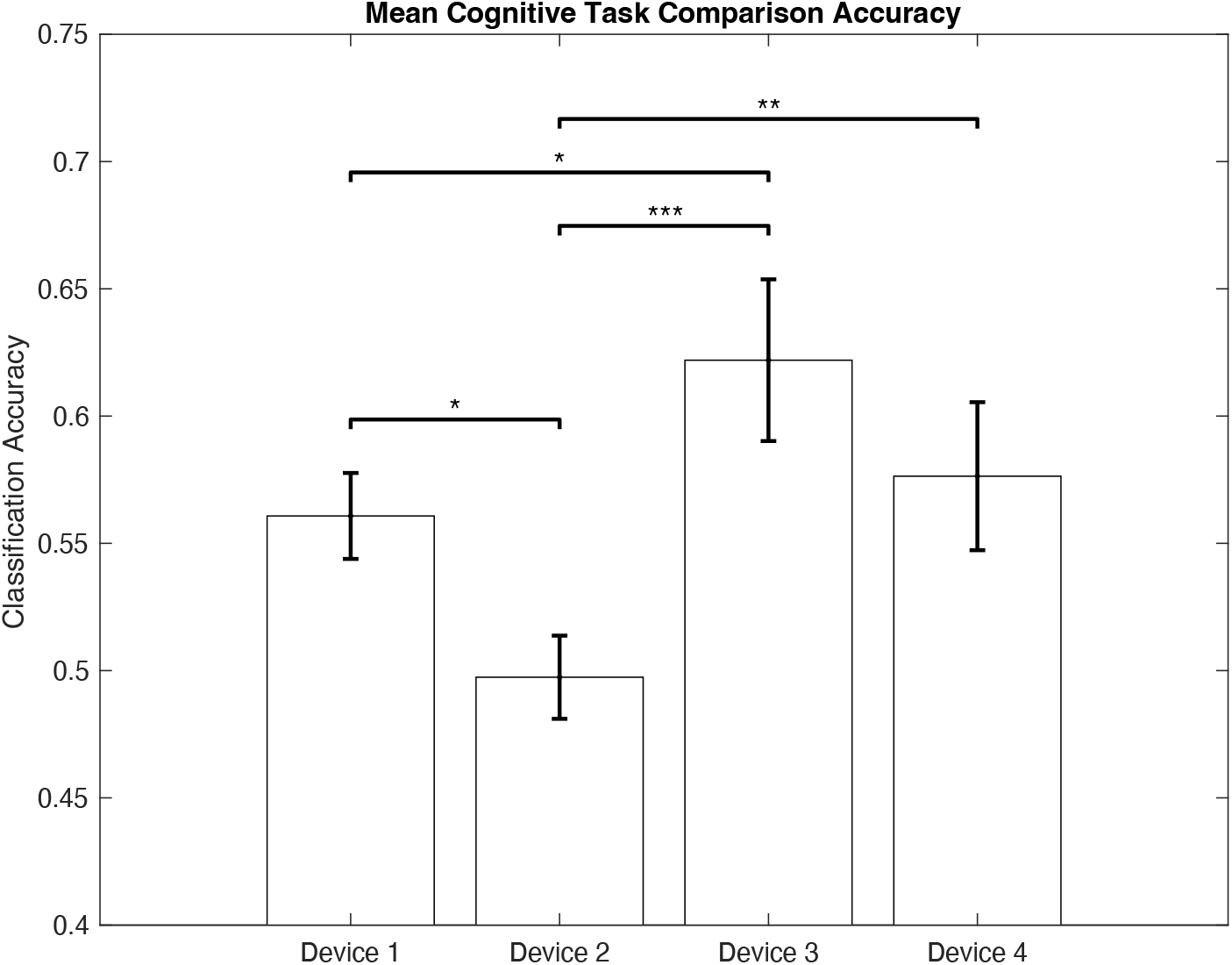
SVM classification accuracy of mean cognitive comparison EEG data, averaged across scans. Error bars display standard error, and brackets display statistical difference as follows: * - p < 0.05, ** - p < 0.01, *** - p < 0.001.

#### 3.2.2 Variations within and between Participants

Classification accuracy in differentiating between cognitive task signals varied both within participants and between participants, as demonstrated in Figure 5. Again, classification accuracies did not differ from participants’ first scan and second scan for Device 1 (p = 0.832), Device 2 (p = 0.961), Device 3 (p = 0.211) or Device 4 (p = 0.134).

**Figure 5.**
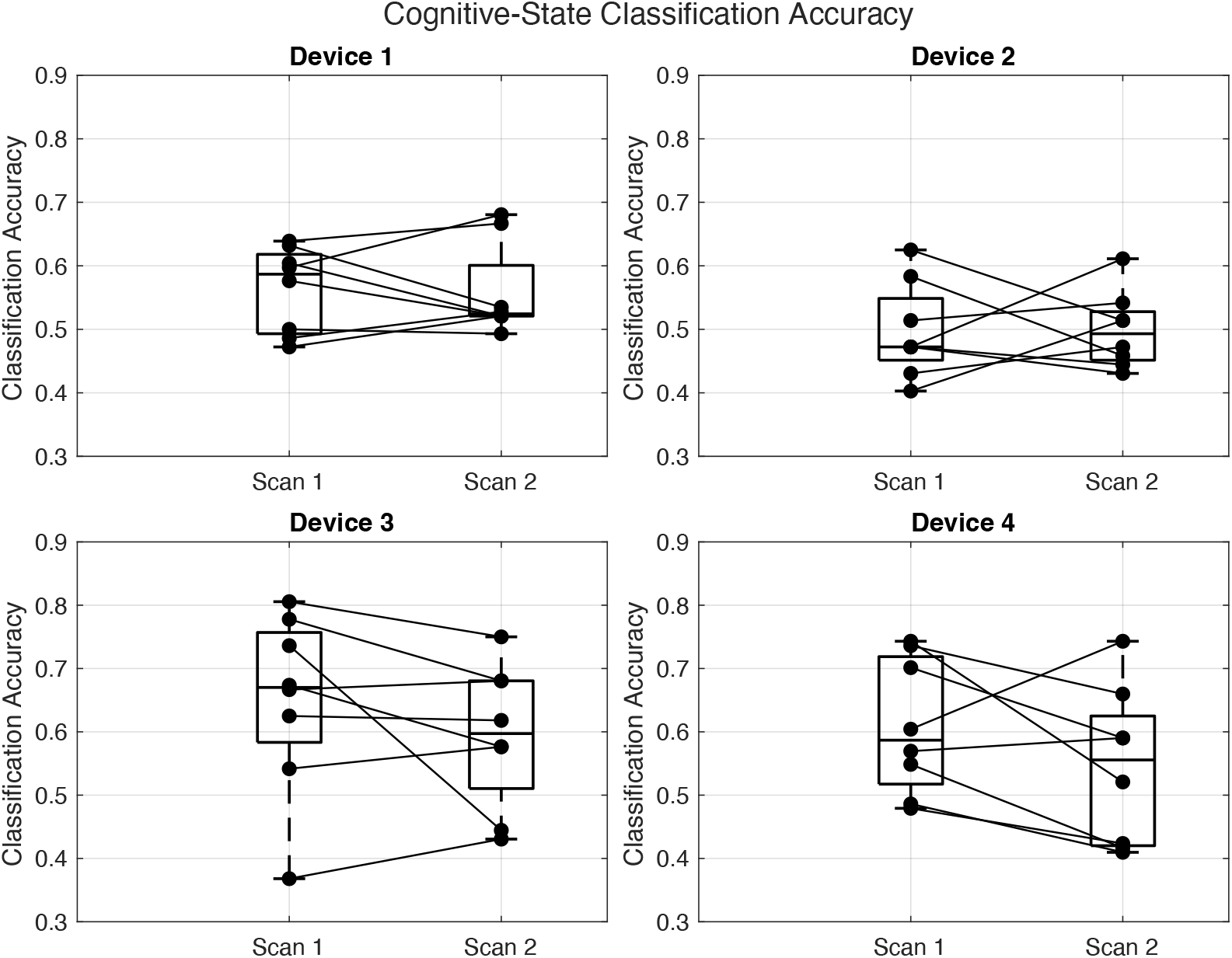
Paired t-test plot of SVM classification accuracy for mean cognitive comparison EEG data by de-vice. Accuracies have been separated out by scan, with classification accuracies corresponding to the same participant linked for each device.

#### 3.2.3 Variations Between Headsets – User-Friendliness

Comparing headsets for user-friendliness, as assessed under the SUS protocol (described in Methods), illustrated differences between headsets (see Figure 6). In terms of average SUS score across the eight participants, Device 1 scored 52.50, Device 2 scored 70.94, Device 3 scored 53.12 and Device 4 scored 79.06. Device had a significant impact on usability score (p = 0.027), encouraging downstream pair-wise device comparisons. These comparisons illustrated Device 4 as more user-friendly than Device 3 (p = 0.014) and Device 1 (p = 0.012). Device 4 and Device 2 were not significantly different (p = 0.421), neither were Device 3 and Device 2 (p = 0.084), Device 3 and Device 1 (p = 0.950) and Device 1 and Device 2 (p = 0.074).

**Figure 6.**
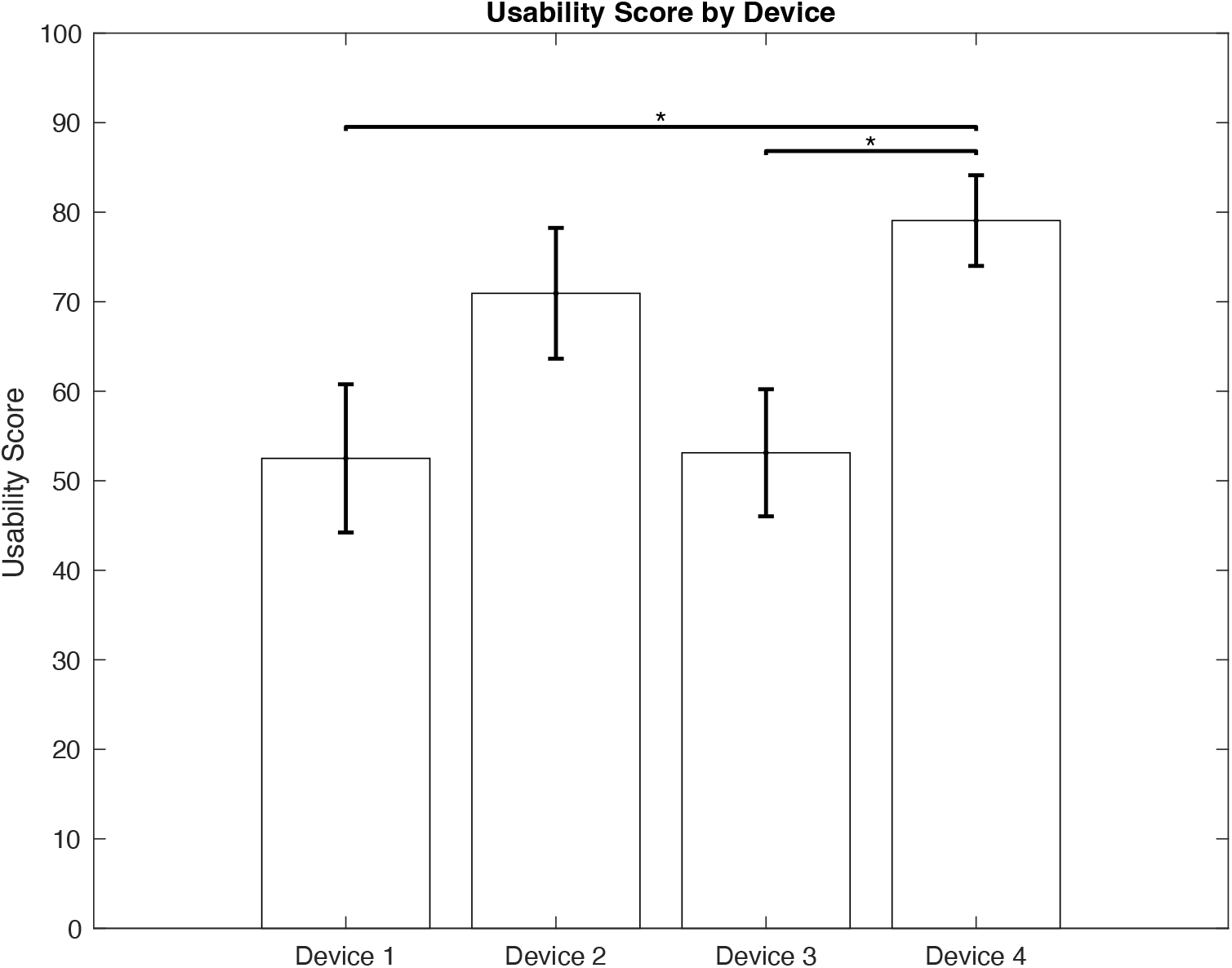
Mean SUS usability scores by headset. Error bars display standard error, and brackets display statistical difference as follows: * - p < 0.05, ** - p < 0.01, *** - p < 0.001.

## 4. Discussion

### 4.1. Comparing Headsets Using Traditional Methods

Testing the four wEEG headsets with this method identified clear differences in resolution (via classification accuracy), with all four headsets performing above chance (i.e. with classification accuracy > 0.5) at differentiating between eyes open and eyes closed signal (as per Figure 2). Under this method of comparison, classification accuracy varied within and between participants, but did not show any inconsistency from participants’ first scans to their second scans (Figure 3). Both the inter- and intra-subject variability would be expected given usual EEG variation between subjects due to age, gender or other factors [78, 79], and within subjects due to concentration levels or other personal factors [80, 81], something that is uniformly seen across all forms of EEG and wEEG.

Type of headset significantly affected classification accuracy, with significant differences noted between Device 2 and all other devices and Device 1 and Device 3 (Figure 2). Under a traditional comparative model, interpreting these results would lead to the conclusion that Device 2 has the lowest resolution, with Device 3 also outperforming Device 1, with a classification accuracy of 0.98 between eyes open and eyes closed states. Device 4 and Device 1 as well as Device 4 and Device 3 were statistically indistinguishable, leaving some ambiguity in technology differences. Potentially relying on this typical analysis could lead to device selection for cognitive monitoring based on very limited performance information.

This insight into resolution derived from traditional analysis roughly follows what we might expect from the four headsets tested. Device 3, Device 1 and Device 4, outperforming Device 2, reflects the resolution that might be expected on the basis of quantity of electrodes (with more electrodes usually being associated with higher resolution [82]), with Device 3 (32), Device 1 (5) and Device 4 (16) all with more electrodes than Device 2 (2) (Table 1). Device 3 also yielding higher resolution than Device 1 is also in line with this theory. That said, given the discrepancy in electrode quantity, it might be expected that Device 3 be superior to Device 4 which in turn should have higher resolution than Device 1, but this is not confirmed by Eyes Open/Eyes Closed comparative analysis.

### 4.2. Comparing Headsets Using Novel Comparative Protocol

Analysis of mean classification accuracy for cognitive task pairs (i.e. across all six cognitive comparisons that were performed) across all participants illustrated differences in headset resolution in being able to specifically and sensitively identify RSN activation associated with functional needs. Three out of the four headsets (all but Device 2) performed better than chance at distinguishing between different cognitive tasks, and therefore different types of cognitive activity (Figure 4). Again, little consistency was seen inter- and intra-subject with all headsets, with no significant differences between participants’ first and second scans (Figure 5). Headset type again was found to have a significant impact on resolution (Figure 4). Pair-wise comparisons demonstrated an identical hierarchy to the benchmark traditional method, with Device 1, Device 3 and Device 4 all supporting better resolution than Device 2, and Device 3 superior to Device 1, but no other significant differences.

While traditional analysis would have relied on this hierarchy to inform headset selection, the new protocol tested here also incorporated assessment of user-friendliness (via SUS scores). In their evaluation of the SUS method, Bangor et al. suggest that usability scores over 70 are considered ‘passable’, while scores below would be considered a sign of serious technology flaws [83]. While this benchmarking is largely arbitrary, it can provide a good understanding of general headset usability and appetite for use.

Under this interpretation, independent headset analysis would illustrate that only Device 2 and Device 4 met the ‘passable’ threshold, highlighting the necessity of this sort of usability assessment within evaluation of hardware, bringing into question any notion of headset quality yielded by analysis of resolution alone. Indeed, comparing headsets on the strength of user-friendliness shows a completely different hierarchy of quality to that offered by resolution outcomes. Analysis shows Device 4 to be more user-friendly than Device 3 or Device 1, but not Device 2, although Device 2 did not meaningfully differ from Device 3 or Device 1 (see Figure 8). Importantly, these results are not consistent with resolution hierarchies alone. This supports the value of this new protocol for more nuanced differentiation between headsets for clinical use cases, with usability outcomes adding to the mosaic of headset differences, significant especially with movement towards at-home, user-led monitoring.

### 4.3. Improving Clinical Decision Making

When we consider the emergence of varied clinical use cases for wEEG technology – ranging from understanding mental health disorders and mental states [84, 85], to sleep disorder analysis [86], to epileptic seizure monitoring [87] – it is clear that each of these uses has different needs from a headset (e.g. sleep disorder analysis might necessitate prioritisation of comfort [88], while mental health or mental state monitoring requires high resolution in picking up specific cognitive patterns [89]). Choice of device should therefore depend on the efficacy and suitability of any given headset for a specific application to ensure that any technology used is fit for purpose. As such, to ensure that clinical decision making is informed to meet these diverse application-specific needs, our analysis of headsets needs to match these evolving needs and provide information that is functionally relevant.

Comparing the picture of headset information offered by traditional methods to that offered by our new protocol, we can see that incorporation of cognitive tasks for assessment of headset resolution, as well as usability analysis, provides a more complete and nuanced understanding of headset differences. Under traditional methods, headset comparisons show Device 1, 3 and 4 outperforming Device 2 and Device 3 also outperforming Device 1. With only this information available, choosing a device becomes a relatively simple decision with Device 3 seeming most suitable for monitoring applications with what would typically be interpreted as the highest EEG resolution. Under the new method proposed here, by assessing cognitive specificity, not only are we able to confirm that headset resolution for targeted applications actually mirrors that suggested by simplified, traditional methods, but we also collect additional information enabling more informed decision-making. Considering usability outcomes would show Device 4 to be more user-friendly than Device 1 or Device 3. Collating this information could lead to the conclusion that for at-home clinical applications that rely on user adoption, Device 4 might actually be a more suitable headset, being statistically indistinct cognitive resolution to Device 3 but more user-friendly. Equally, for applications that require high specificity in detection of cognitive activity, Device 3 could be preferred with mean classification accuracy between task-induced states of greater than 0.6. The conclusions afforded by this proposed method are not only more nuanced and flexible, but also more relevant to the use of wEEG technology within newer clinical applications that could require both specificity in detecting cognitive activity and comfort for at-home use.

Under this new layered analysis, irrespective of whether user needs necessitate comfort, cognitive specificity or both, headset selection can be more informed than under traditional methods that unjustifiably prioritise arbitrary markers of resolution with little to no relevance to these new demands. As such, for the variety of clinical use cases that wEEG can increasingly help address, but especially for at-home use in the monitoring of cognitive activity, this new comparative protocol represents better functional approximation of headset performance. By designing a method of headset evaluation that can better inform users on headset efficacy related to desired headset function, we can therefore facilitate more scientifically sound selections of wEEG headsets for clinical purposes, in a field with a broad spectrum of technology structure and quality.

### 4.4. Limitations and Future Directions

While the new protocol proposed here is doubtlessly an improvement on common methods of comparing wEEG technology, it would be reductive to suggest that it is a flawless method on the strength of its demonstration in this study.

In addition, while the cognitive paradigms chosen here to make up the protocol are founded in the literature, these types of task-based paradigms are not necessarily well-established in past EEG research. The tasks selected here were chosen with respect to the clinical aims of Resonait Medical Technologies in using wEEG technology for the support of mental health conditions. As such, these tasks, while still very much grounded in clinical relevance, may not canvas all possible applications of wEEG technology – e.g. sleep monitoring might prioritise detection of sleep rhythms that are not necessarily linked to any specific network activation [88]. For this method to have the potential for standardised, universal assessment of wEEG technology, both the broader clinical significance of the functional networks selected, as well as the efficacy of the chosen tasks to elicit functional network activation need to be validated. The former will become easier with the validation of clinical use of wEEG systems over time, as wEEG headsets improve in quality and become more commonplace within clinical and commercial practice, while the latter would be possible with further experimentation – e.g. by performing these tasks with traditional clinical-grade EEG, or even functional Magnetic Resonance Imaging (fMRI), to ensure that local brain activity matches what would be expected for activity in the functional network ‘linked’ to each task [91]. Importantly, carefully selecting tasks of significance has the additional potential for downstream analysis into specific task-pair classification outcomes. Indeed, part of the value of this new proposed protocol is the potential for increased specificity in elucidating functional aptitude of wEEG technology in an area where new functions are constantly being identified. As such, further enquiry into individual classification accuracies (rather than general cognitive resolution) could give us insight into how headsets vary in their ability to specifically detect activity in some RSNs over others, something that can be of clinical relevance. For example, for neural monitoring in the treatment of depression, DMN activity is of specific interest [92, 93]. Accordingly, when choosing wEEG technology for this purpose, headset ability to discriminate DMN activity from other functional networks is both relevant and important. Future consideration should definitely be given in the selection of cognitive tasks for headset comparison to maximise the value of the data yielded.

The large inter- and intra-subject variability seen throughout this work, while fairly consistent with wEEG technology through the literature [97, 98], may additionally limit the findings presented here. In terms of inter-subject variability, given the low data volume, the spread of results may not necessarily be typical of a larger cohort of participants. Beyond the low data volume, the participants of this study were also not representative of wider populations. Sex was not distributed evenly, a potential confounding factor as sex differences have been shown to cause systematic differences in EEG signal [99-101], possibly contributing to the inter-subject variability in pattern classification seen in this pilot. Age in this trial was not very widely distributed either, with age also influential in the strength of and patterns within EEG signal<u>s</u> [102, 103], thus possibly limiting the generalisability of the headset performances on this set of participants. Demographic differences impacting EEG are not limited to age and sex, with other factors, even including hair type and texture, able to affect data quality [104]. Continuing this experimentation with a larger, diverse set of participants would minimise any potential skews and be extremely informative on the generalisability of the results achieved on the small cohort examined here, ensuring the statistical validity of results is consistent with more data.

Finally, in terms of intra-subject variability, it is possible the learning of behaviours as more scans were performed was a driver of these differences. By the time participants reach their second scan with any given headset, they would have performed all cognitive tasks at least 24 times (six times per scan, and all four headsets were used at least once before they were used for a second scan). Skill acquisition from repeat task completion has been shown to increase EEG strength in the past [105], meaning that the EEG patterns in participant’s second scans with each device may differ from their first scan, resulting in intra-subject discrepancies in classification accuracy. This effect was addressed as much as possible with randomisation of participant scan order, with the aim of balancing across the headsets, but it is difficult to completely mitigate. While this is a potential confounding factor, it should be noted that despite any individual intra-participant differences in classification accuracy, as demonstrated in Figure 3 and Figure 5, there were no significant changes across the cohort from Scan 1 to Scan 2 for any of the tested devices. Further investigation into participants who experienced significant changes between scans with any given device could provide increased insight and better inform any future evolutions of this protocol.

Moving forward, for this protocol to see eventual standardisation and broader uptake across the industry, more thorough investigation would be needed to address the drawbacks in this experimentation. Future work should seek to repeat this experimentation with a bigger and more diverse sample size of participants to further support the generalisability of the results presented here, and therefore the value of this comparative protocol. Additionally, future investigation could look to substitute out or supplement the cognitive tasks chosen here with other tasks that are functionally linked to various other clinical applications to improve universal applicability of this new method. If these future steps continue to show promising results, this protocol has the potential for broader standardisation in the analysis of wEEG technology, something that would be incredibly significant for an industry marked by inconsistency preventing meaningful impacts to patient care.

## 5. Conclusion

With the traditional methods of comparing wEEG technology increasingly limited in their ability to provide meaningful insight into headset suitability for emerging clinical applications and at-home environments, this research offers the beginning of a solution to an unaddressed problem. By leveraging MVPA methods and standardised usability testing, the protocol designed and tested here is able to maintain the detail provided by well-established methods, while also enabling possibilities for function-specific and user-specific analysis, ultimately making progress towards ensuring that selection of a wEEG device for novel clinical applications through a data-driven way. By providing better guidance to make these decisions, the work performed here can hence assist in ensuring that the new pathways that wEEG can unlock are not lost or compromised through outdated methods of evaluation and verification.

## Data Availability

All data used for this work will not made publically available, beyond what is already contained in the manuscript.

## Notes

### Competing Interest Statement

Cameron Higgins is an employee, shareholder and Director of Resonait Medical Technologies Pty Ltd. The other authors declare no competing interests.

### Author Declarations

The ResearchIntegrity and EthicsAdministrationof The University of Sydney gave ethical approval for this work (2024/HE001864).

### Summary of Updates

Updated competing interest statement and author affiliations.

